# Extending the OMOP Common Data Model to Support Observational Peripheral Vascular Disease Research

**DOI:** 10.64898/2026.02.01.26345276

**Authors:** Peter J. Leese, Tomas McIntee, Sydney E. Browder, Mirjami Laivuori, Olamide Alabi, Katharine L. McGinigle

## Abstract

**Background:** Peripheral artery disease (PAD) and chronic limb-threatening ischemia (CLTI) cause substantial morbidity and mortality, yet research progress is limited by fragmented, non-standardized data. The Observational Medical Outcomes Partnership (OMOP) Common Data Model (CDM) provides a standardized framework for electronic health record (EHR) research but lacks domain-specific detail for peripheral vascular diseases. This study aimed to develop and test a vascular-specific OMOP CDM extension to improve data standardization, enable reproducible real-world analyses, and support precision medicine research in PAD and CLTI.

**Methods:** We identified patients with PAD, CLTI, or diabetic foot ulcers who sought care within the UNC Health System between April 2014 and July 2024. Standard OMOP tables were supplemented with peripheral vascular laboratory (PVL) data and state death records. Intermediate tables were designed for key clinical domains (e.g., smoking, comorbidities, revascularizations) to enhance reusability. Predictive models for revascularization and mortality were developed using logistic regression with Bayesian weighting and Markov Chain Monte Carlo feature selection.

**Clinical Application:** The revascularization model displayed high performance with and without important vascular variables (AUC = 0.970 and AUC 0.969, respectively), while the mortality model demonstrated moderate accuracy (AUC = 0.656) that improved with inclusion of vascular-specific features (AUC = 0.752).

**Conclusions:** This vascular OMOP extension represents one of the first specialty-specific frameworks for peripheral vascular research. By extending the OMOP CDM to a vascular domain, this work advances both the technical framework and scientific capability of real-world data research in limb preservation and precision vascular medicine.

## Introduction

Peripheral artery disease (PAD) is a chronic, occlusive arterial condition of the lower extremities that imposes substantial limb-based morbidity and mortality from cardiovascular causes.^1,2^ Its clinical presentation is widely variable, shaped by comorbidities such as hypertension, diabetes, hyperlipidemia, and smoking, as well as social and environmental factors. The arterial plaque burden, ischemic tissue injury, and coexisting illnesses complicate treatment decisions and contribute to the risk of limb loss.^3,4^ Individuals without PAD, but with microvascular disease and diabetic foot ulcers, also share many of these presenting characteristics, risk of amputation, and poor survival rates.^5^ The umbrella clinical term for these chronic conditions that put a patient at risk for limb loss is called chronic limb-threatening ischemia (CLTI).^3^ To treat CLTI, clinicians must balance life-preserving medical goals with limb-based interventions over the course of the disease; however, heterogeneous presentation, multiple medical and surgical therapeutic options, and differential patient responses make optimal long-term management challenging.^6^

Despite its prevalence and clinical importance, PAD and CLTI research suffers from a weak evidence base.^7^ Few randomized controlled trials have focused on PAD and large, prospective cardiovascular cohorts lack peripheral vascular-specific variables.^8^ Existing PAD datasets are typically small, single-institution efforts requiring intensive manual abstraction. Electronic health records (EHRs) and their rich, detailed longitudinal data provide an opportunity to improve and standardize datasets to efficiently and impactfully conduct large, multi-institutional studies.^9^

The Observational Medical Outcomes Partnership (OMOP) Common Data Model (CDM) is a widely adopted data architecture used to standardize electronic health data.^10^ The OMOP CDM facilitates reproducible observational research through a standardized vocabulary and a relational schema formally linking more specific and more general terms, which serve as a framework to organize data. Multiple NIH-funded programs now use the OMOP CDM to store and standardize clinical and genomic data.^11,12^ Additionally, UK Biobank data has been transformed into the OMOP CDM for large-scale research on COVID-19.^13^ As the utilization of real-world data research has grown, and use of the OMOP CDM with it, there have been many advances in EHR-based studies. For example, one study utilized the OMOP CDM across 47 data partners in Rhode Island to standardize health information exchange data in order to calculate longitudinal atherosclerotic cardiovascular disease risk on more than 60,000 individuals, while another study analyzed treatment pathways for hypertension, type II diabetes, and depression from 2005 to 2015.^14,15^

Although OMOP is useful for broad EHR research, the baseline OMOP model lacks specialty-specific data necessary for more specialized research and nuanced clinical phenotyping.^16–18^ The absence of domain-relevant constructs in the base OMOP CDM has prompted the development of specialty-specific extensions, such as the OMOP Oncology Extension and Radiology CDM.^19,20^ These previous efforts demonstrated that specialty-specific extensions can successfully standardize specialty data while retaining OMOP’s broader interoperability benefits. Given the complexity and unique terminology of PAD and CLTI—including arterial anatomy and occlusive plaque location, severity of the perfusion deficit, wound location and classification, and open surgical and endovascular interventions—we developed a peripheral vascular-focused OMOP extension to support large-scale real-world data analysis and precision medicine applications in PAD and limb preservation.

## Materials and Methods

### Data Source and base OMOP CDM

Our institution maintains a robust healthcare data and informatics ecosystem designed and managed by the North Carolina Translational and Clinical Sciences (TraCS) Institute within the University of North Carolina at Chapel Hill School of Medicine. Much of this data is generated by clinical care from the UNC Health System in the EPIC EHR application and is stored in the various EPIC databases, including Clarity for reporting. For research purposes, data from Clarity is transformed into the PCORnet and OMOP CDM. The TraCS Institute is active in OMOP CDM and vocabulary work in both EHR and claims data sources across both single-site instances and large, harmonized multi-site projects.^21–24^

### Vascular Cohort Identification

In this foundational work of building common, harmonizable data architecture for peripheral limb-threatening conditions, we opted for a registry design, including only patients with a diagnosis that could lead to limb threat and not negative controls. A formally validated computable phenotype with test statistics using ICD10 or SNOMED codes for PAD, diabetic foot ulcer, or CLTI was not identified in phenotype registries nor in medical literature at large.^25^ As a result, we reviewed a variety of literature that developed relevant PAD identification algorithms, reviewed data dictionaries from national vascular registries, and consulted with clinical vascular specialists at multiple institutions.^26–30^ Based on these reviews and consultations, we relied on condition occurrence source values in OMOP to identify highly likely PAD and CLTI patients. The logic, terminologies, and codes used to identify this cohort are illustrated in the supplement. Patients who sought care within the UNC Health System (inpatient or outpatient) were identified between April 2014 and July 2024.

### Basic Datamart Architecture

After identifying likely PAD and CLTI patients, we extracted all OMOP data for these patients from the core OMOP tables. These data included demographics, diagnoses, procedures, medications, labs, encounters, vital status, and patient reported data from the respective OMOP tables (person, condition occurrence, procedure occurrence, drug exposure, measurements, visit occurrence, death, and observations). Beginning with the OMOP warehouse of all EHR data, a derived datamart was designed to support the PAD registry development. For the datamart these tables were maintained in their OMOP relational format and then supplemented with peripheral vascular laboratory (PVL) measurements from the EPIC Clarity database. These PVL measurements were brought in as an ‘OMOP sidecar’ so that while they could be linked to the OMOP data, they were not mapped to the OMOP standard. The architecture of the datamart is illustrated in Figure 1.

**Figure 1.**
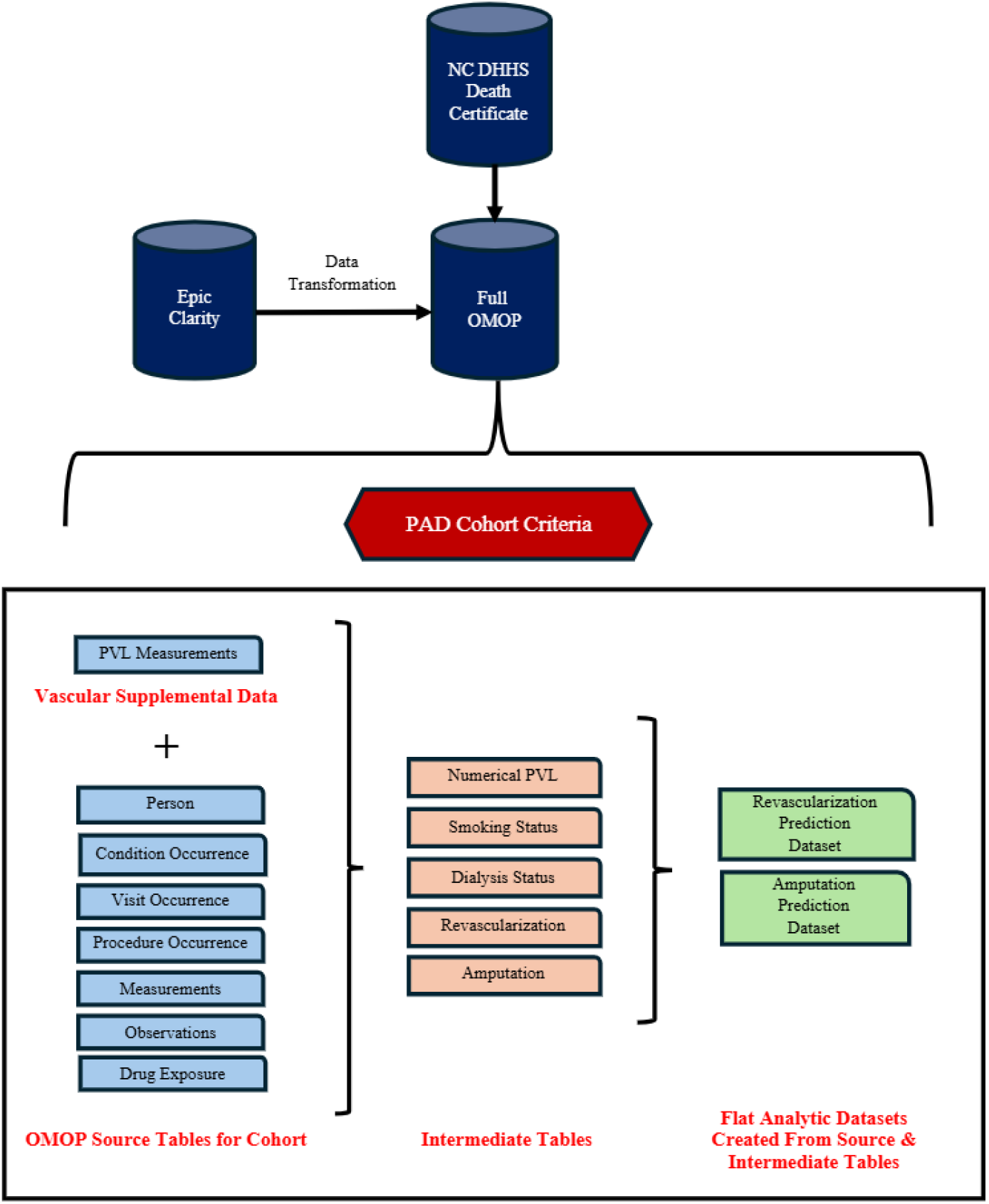
OMOP Vascular Extension Data Architecture

### Vascular Supplemental Data

Due to the aforementioned gaps in OMOP for vascular research, it is necessary to supplement standard OMOP EHR data with additional vascular-relevant data. After consideration of various vascular-relevant clinical data features, the most important supplemental data identified to confirm PAD diagnosis and severity was PVL measurements like the ankle-brachial index and toe pressure.^3^ Additionally, due to the importance of patient vital status as a long-term outcome, EHR patient mortality was supplemented with data from UNC’s state-level death certificate linkage.^31^ The PVL measurements were obtained directly from EPIC Clarity from the text of arterial duplex ultrasound reports. At our institution, the data from the PVL are entered in a free-text format rather than in discrete data fields within EPIC. As such, numeric PVL values were extracted algorithmically from the report text via structured query language (SQL). As text, these data are challenging to work with and we developed code to identify as many discrete PVL measurements as possible; however, we anticipate enhancing PVL extraction from text reports via large language model.

### Intermediate and Derived Tables for Extensibility

An intentional feature of the vascular OMOP extension is the derived, hierarchical relational model to facilitate reusability and extensibility across studies. At the top level, the cohort is identified and the ‘raw’, source OMOP data for these patients is pulled – this re-creates an OMOP database for our PAD patients and this data is then supplemented with the ‘raw’ PVL text data and external death data. This is a very broad dataset that can facilitate many different activities: retrospective analyses across diverse focus areas, broad population level assessments, and patient identification for recruitment activities. To facilitate rapid development in frequently used areas, we design and build a layer of intermediate tables derived from the cohort OMOP tables for common feature spaces. For instance, smoking is relevant to almost any subsequent recruitment or analysis activity for PAD; however, smoking can be identified (such as diagnosis vs patient reported data) and defined (binary, pack years, etc) in myriad ways which is then typically built out very specifically and narrowly for a specific research application. Instead, in our PAD datamart we designed and built an intermediate table that assembles all smoking relevant data into a ‘smoking status’ intermediate table derived from all core OMOP tables such that this table can then be used to define smoking for any subsequent analysis in any way without needing to re-assess and analyze the vast and enormous full OMOP tables for each study. In this way, we attempted to build these intermediate tables to capture the entire feature-space such that any specific smoking feature can be defined purely from the ‘smoking status’ intermediate table. This was then repeated for all identified, relevant feature spaces for PAD, such as diabetes, dialysis dependence, etc. Currently in our initial development of the OMOP PAD datamart, we have intermediate tables for smoking, amputations, revascularizations, dialysis dependence, and Charlson Comorbidity Index (CCI). Due to known lack of validation and methods for calculating CCI in both EHR data and OMOP we relied on our own algorithm, largely based on Quan’s 2005 version.^32–38^

### Patient Summary Table – Analytic Table Design

Focused analytical tables or flat-file datasets for focused research purposes were created utilizing the raw OMOP tables and the intermediate tables to create wide format aggregated to the relevant unit (patient vs episode, etc) with relevant features for analysis activities. The concept of the PAD OMOP datamart is that once the intermediate layer of tables is derived from the raw tables, it is possible to more efficiently design and build flat, focused patient summary tables for specific research questions.

For studying the precision medicine question of whether or not a patient can be expected to show improved outcomes with a revascularization event, we used a wide summary table that included age, sex, smoking, the cohort entry and exit dates, an operation decision date, drug exposures and key measurements, like lab values indicating severity of comorbidities, and depression status from before the decision date, and limb loss and death outcomes after the decision date. (The operation decision date was a revascularization operation for treated patients and a synthetically generated decision date drawn from the posterior distribution of measured time-to-treat for untreated patients.) This table included one row per patient with key relevant variables, including demographics, death dates (where applicable), cohort entry and exit dates, dates and types of operation for revascularizations and amputations, descriptive statistics for key measurements (a1c, LPA, GFR, LDL cholesterol, and albumin) and prescriptions (statins, antithrombotics, and drugs associated with diabetes).

### Tools, Reproducibility, and Code Availability

Initial build of this OMOP PAD datamart was performed in Azure Databricks and employed a combination of Spark SQL, Pyspark, Python, and R. Subsequently, we have converted this codebase into pure R, SQL, and Python for implementation in non-cloud and non-Spark environments. The analytic pipeline was version-controlled using Git and executed within a secure institutional environment. Data engineering of the peripheral OMOP datamart prioritized FAIR principles of findable, accessible, interoperable, and reusable. While the EHR data in our datamart cannot be publicly shared, all code will be made publicly available. This code is available at: https://github.com/NCTraCSIDSci/nctracs_vascular_foundation/

## Clinical Application of Datamart to Prediction

### Clinical Vignettes

To test the datamart, we developed two analytic applications that we pursued using these data: prediction of revascularization and prediction of mortality in PAD patients. Utilizing the full OMOP source tables and intermediate tables, a patient summary table was developed that included relevant patient and disease features including revascularization and mortality. In addition to literature and clinical informed variable selection for modeling, we also employed large-scale dimensionality reduction focused on approximately 12,000 distinct conditions and observations to identify unique features.

### Feature Selection

The initial feature selection process began with extracting data from the OMOP condition occurrence and observation tables, identifying over 12,000 distinct conditions and observations. To reduce dimensionality, rare conditions and observations (those seen in fewer than 1,800 patients) were mapped to more general ancestor concepts using the OMOP concept ancestor table, limiting rollup to within four levels of ancestry. This initial rollup yielded 1,290 candidate features. A second consolidation step compared the predictive performance of ancestor concepts versus their descendants using logistic regression models for death as an outcome, applying a threshold for acceptable quality of fit loss (0.001). If the outcome did not exceed this threshold, the descendant concepts were further rolled up. After this two-stage rollup process, 1,196 OMOP features remained. To further refine the feature set, domain experts manually grouped 53 related diagnoses into six clinical categories for a final pool of 1,149 OMOP features. We then added 127 vascular features (e.g., demographics and vascular measurements), resulting in a final pool of 1,276 usable features (Figure 2).

**Figure 2.**
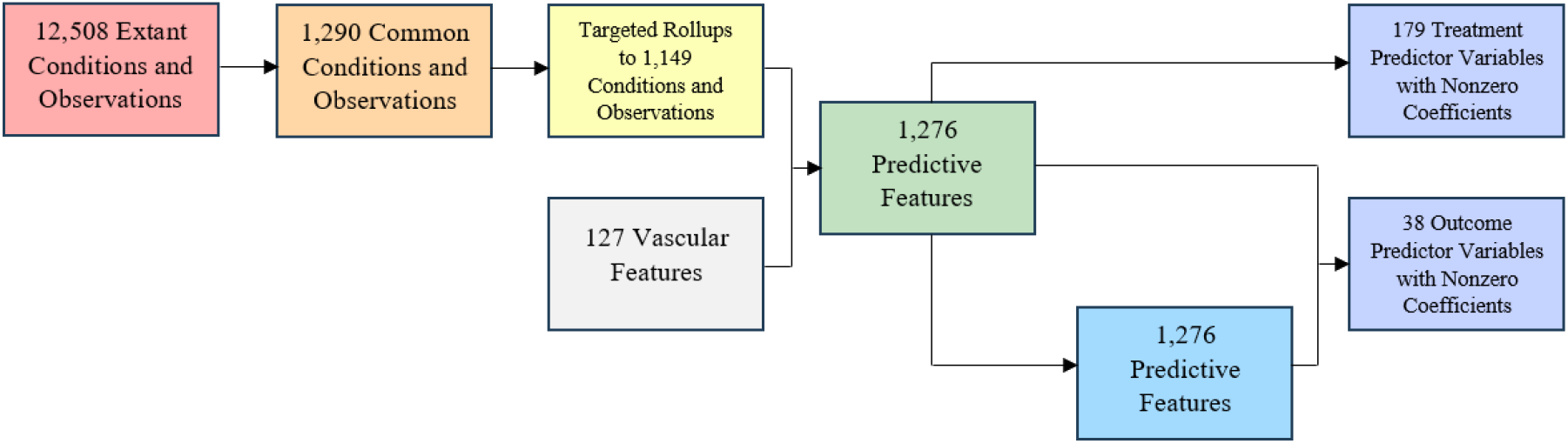
Feature Selection for Case Example

### Building the Full Model

The dataset was divided into a training set of 100,000 patients and a validation set of 70,590. A two-level logistic regression model was constructed for death as the outcome, starting with a prediction of revascularization, followed by an outcome model conditional on revascularization status. Patients without revascularization were assigned a synthetic decision date to align time windows between treated and untreated groups. Inverse probability of treatment weighting (IPTW) was applied using propensity scores to mitigate confounding in the outcome models. Initial feature weighting was based on Bayesian odds ratios, and weights were refined using a Markov Chain Monte Carlo (MCMC) method with repeated sampling and model fitting via glmnet. This process served both as a dimension reduction and variable selection strategy. The final logistic regression model was built using the 200 most consistently important features.

### Model evaluation

The final models were evaluated using a receiver operator characteristic curve, taking the measurement of area under the curve (AUC) as an estimate of overall model quality. AUC for predictive models generally ranges from 0.5 (random assignment of scores) to 1.0 (perfect classification). Two different model variations were considered, one using only the native OMOP variables and the other using the added vascular variables. Performance was evaluated for two models: a final treatment (revascularization) model and a death prediction model. With 1,000 iterations of the MCMC cycle, the final treatment model had exceptionally high performance (AUC = 0.970), even when the highly predictive vascular variables were omitted (AUC = 0.969). This implies that revascularization treatments occur on a highly non-random basis, with revascularizations generally targeting patients strongly in need of revascularization. The death prediction model had considerably lower performance with 1,000 MCMC iterations (AUC = 0.752, OMOP-only AUC 0.656) and did not significantly improve with 2,000 MCMC iterations (AUC = 0.749 & AUC = 0.688), suggesting that this was not a convergence issue. This implies that the OMOP plus vascular extension model provided moderate accuracy in predicting death and this model was able to predict death more accurately than the OMOP-only model. Potential explanations for the diminished performance of the death model are that the most important variables in the model were less independently informative and that death proceeds from numerous factors, making each factor individually weaker for prediction.

## Discussion

To our knowledge, this study represents one of the first vascular-specific extensions of the OMOP CDM. By creating a standardized vocabulary and relational schema tailored to peripheral vascular disease, we extend the OMOP framework into a procedural and hemodynamic specialty domain. The vascular extension supplements core OMOP EHR data with external PVL and external mortality data, demonstrating a generalizable strategy for incorporating specialty-specific information that is not yet represented within the base OMOP model. This work advances the OMOP data architecture while preserving its interoperability, structure, and analytic consistency.

The development of the OMOP vascular extension was motivated by two central challenges in vascular research. First, the absence of a standardized PAD-specific EHR data model has required investigators to create ad hoc variable definitions that impede reproducibility and comparability across studies. Second, the extensive and heterogeneous nature of vascular data— spanning anatomy, physiology, procedure, and outcome domains—creates methodological complexity in identifying the most informative features for research and clinical modeling. Our extension addresses both challenges by introducing harmonized, reusable variable definitions that can be implemented across institutions, and by identifying consistently important vascular features that improve model interpretability and support precision medicine research. In doing so, this work parallels and extends prior OMOP specialty initiatives such as the Oncology and Radiology CDMs, establishing a framework for integrating vascular-specific diagnostic and procedural data into the broader OMOP ecosystem.^19,20^

Model evaluation demonstrated that our revascularization (treatment) prediction model performed with high accuracy both with and without vascular-specific variables, while the mortality model achieved moderate performance, with improvement when the vascular extension was included. The near-equivalent performance of treatment models across both variable sets suggests that procedural decision-making for revascularization is already well-captured in structured EHR data. In contrast, mortality prediction benefited from inclusion of disease-specific features, indicating that domain extensions may yield greater incremental value for multifactorial outcomes such as death, which depend on heterogeneous physiological and procedural factors. These findings underscore the potential for OMOP extensions to enhance outcome prediction and support nuanced, disease-specific modeling within an interoperable framework.

While feasibility studies have been reported for other OMOP extensions, few have used extension data to address complex clinical questions.^39^ In theory, the specialty-specific vocabularies and schemas developed through such extensions can enable analyses that were previously infeasible due to data fragmentation or limited sample size. Broader adoption of disease-specific OMOP extensions—such as this peripheral vascular model—could help close long-standing evidence gaps in real-world clinical research, facilitating multi-institutional studies and accelerating methodologic standardization. As with prior extensions, achieving scalability will depend on alignment with OHDSI governance processes, harmonized vocabularies, and local OMOP familiarity and data engineering capabilities across health systems.

Our work has several limitations. First, vascular data integrated from PVL reports was limited to ankle–brachial index and toe pressure measurements; additional data such as computed tomography angiography (CTA) or magnetic resonance angiography (MRA) could provide a more complete representation of vascular anatomy but remain difficult to integrate within the current OMOP structure. Second, our implementation was single-institutional and focused on a core subset of vascular variables. Further validation across institutions and data environments will be necessary to ensure portability and robustness. Lastly, while our modeling demonstrated promising performance, omitted EHR variables and reliance on structured data sources may have limited the full potential of predictive accuracy.

Future work will expand the vascular extension to include imaging, procedural, and hemodynamic data using standardized OMOP Measurement and Observation constructs. Cross-institutional validation via OHDSI collaboration and incorporation of advanced text and image extraction methods—such as large language models for PVL reports—represent key next steps. Ultimately, this peripheral vascular OMOP extension establishes a foundation for reproducible, large-scale, and longitudinal analyses of peripheral artery disease and chronic limb-threatening ischemia.

### Conclusion

Accessing the full capabilities of EHR data has vast implications for disease-specific health research. Our peripheral vascular OMOP extension adds disease-specific vocabulary to the increasingly popular OMOP CDM. This extension is built to allow health professionals to answer numerous research questions that remain unanswered due to a paucity of large datasets containing extensive, multi-institution, longitudinal data related to the diagnosis and treatment of peripheral vascular disease. By extending the OMOP CDM to a vascular domain, this work advances both the technical framework and scientific capability of real-world data research in limb preservation and precision vascular medicine.

## Data Availability

Data produced in the present study are not available.

## Author Contributions

**Peter Leese:** Conceptualization, Data curation, Formal analysis, Investigation, Methodology, Project administration, Software, Supervision, Validation, Visualization, Writing-Original draft preparation, Writing – Reviewing and Editing. **Thomas McIntee:** Data curation, Formal analysis, Investigation, Methodology, Software, Validation, Visualization, Writing-Original draft preparation, Writing – Reviewing and Editing. **Sydney Browder:** Project administration, Visualization, Writing-Original draft preparation, Writing – Reviewing and Editing. **Mirjami Laivuori:** Visualization, Writing-Original draft preparation, Writing – Reviewing and Editing. **Olamide Alabi:** Conceptualization, Writing – Reviewing and Editing. **Katharine McGinigle:** Conceptualization, Funding acquisition, Project administration, Supervision, Visualization, Writing-Original draft preparation, Writing – Reviewing and Editing.

