## Supplement for "Extending the OMOP Common Data Model to Support Observational Peripheral Vascular Disease Research"

Codes used to identify patients with PAD or at-risk of developing PAD

| Included / Excluded | Granularity | Terminology | Codes |
|---|---|---|---|
| Include | exact | ICD10 | I73.89, I73.9, I77.70, I77.72, I77.76, I77.77, I77.79, I79.8 |
| Include | 5 digit or more specific | ICD10 | E08.5, E09.5, E10.5, E11.5, E13.5 |
| Include | 3 digit or more specific | ICD10 | I70, I74, I75, I76, I96, L97 |
| Exclude | 3 digit or more specific | ICD10 | I71, I72 |
| Exclude | exact | ICD10 | I70.1 |

Codes used to identify relevant procedures

| Procedure | Terminology | Codes |
|---|---|---|
| Amputation | SNOMED | 4345680, 42536891, 4176170, 4308713, 4103642, 4202322, 133088, 135722, 196613, 4101660, 4103640, 4103638, 4113102, 2105806, 4272232, 4338257, 4195136, 4302020, 4143797, 2105450, 4159766, 2101638, 2105448, 2105223, 2006243, 2105449, 4054983, 4143795, 4264289, 2105211, 4078404, 2105222, 40480578, 2105447, 2105451, 2006242, 36675618 |
| Revascularization | CPT | 34201, 34203, 35302, 35303, 35304, 35305, 35306, 35331, 35351, 35355, 35361, 35363, 35371, 35372, 35537, 35637, 35538, 35638, 35539, 35647, 35540, 35646, 35556, 35656, 35558, 35661, 35563, 35663, 35565, 35665, 35566, 35666, 35570, 35571, 35671, 35583, 35585, 35587, 35621, 35623, 35654, 35700, 35875, 35879, 35881, 35883, 35884 |
| Revascularization | ICD10 | 04CK0ZZ, 04CL0ZZ, 35656, 35371, 34201, 35666, 047L3ZZ, 04CL3ZZ, 041L0JL, 35661, 047K3ZZ, 04CK3ZZ, 35372, 041K0JL, 047Q3ZZ, 35302, 047P3ZZ, 34203, 047N3ZZ, 35875, 047R3ZZ, 047U3ZZ, 047M3ZZ, 047T3ZZ, 047S3ZZ, 04CM0ZZ, 35700, 04CN0ZZ, 35556, 35646, 04CM3ZZ, 047C3DZ, 047D3DZ, 04CN3ZZ, 047K3DZ, 047L3DZ, 047H3DZ, 35876, 35566, 047J3DZ, 047D3ZZ, |

| | | |
|---|---|---|
| | | 047C3ZZ, 047Y3ZZ, 04CH0ZZ, 041L09L, 04CJ0ZZ, 047J3ZZ, 35654, 047L34Z, 35583, 04CY0ZZ, 047H3ZZ, 041K09L, 041K0JJ, 04100JK, 35585, 04CP3ZZ, 047K34Z, 04CR3ZZ, 04CS0ZZ, 041L0JH, 04CC0ZZ, 35571, 04CR0ZZ, 041L0JN, 04CU3ZZ, 04CP0ZZ, 047N34Z, 047M34Z, 04CQ3ZZ, 04CT3ZZ, 04CS3ZZ, 04CD0ZZ, 04CQ0ZZ, 047M3DZ, 35351, 047N3DZ, 35303, 04703DZ, 04CU0ZZ, 35883, 041K0JN, 047K0ZZ, 35665, 047D34Z, 041K09N, 04CT0ZZ, 047H0DZ, 067D3DZ, 047C34Z, 04C00ZZ, 35621, 041K0ZL, 047L0ZZ, 047C0DZ, 047P34Z, 04CH3ZZ, 047J34Z, 047D0DZ, 047H34Z, 047Q34Z, 041L09N, 047J0DZ, 03743DZ, 047S34Z, 041K0JM, 04CJ3ZZ, 04CD3ZZ, 04CC3ZZ, 041K0JQ, 041L0JM, 35671, 04CY3ZZ, 041N09Q, 041K0JH, 047P3DZ, 041L0KL, 03150J8, 35638, 047U34Z, 047K0DZ, 041L0ZL, 047T34Z, 041K0KL, 047V3ZZ, 041L0JJ, 047Y3DZ, 35304, 047H0ZZ, 041K0KN, 041L09M, 03160J7, 041M09Q, 057Y3DZ, 03150J6, 047N0ZZ, 35331, 041L0KN, 047P0ZZ, 35305, 047R34Z, 047M0ZZ, 04BL0ZZ, 047C0ZZ, 04BK0ZZ, 04100J8, 047W3ZZ, 041L0ZH, 047C04Z, 35558, 067F3DZ, 35587, 047Y34Z, 047D0ZZ, 03150J9, 047K04Z, 041K09M, 04LL3DZ, 047H04Z, 35355, 047L0DZ, 041L09Q, 041K0ZJ, 03160JC, 047T3DZ, 047J04Z, 047J0ZZ, 047L04Z, 041K09Q, 041K0JK, 04LK3ZZ, 047S3DZ, 04703ZZ, 047S0ZZ, 04LK0ZZ, 041L0ZN, 04104J8, 047E3ZZ, 04100JJ, 35647, 35623, 04100ZK, 047M0DZ, 047R0ZZ, 04CK4ZZ, 047D04Z, 04C03ZZ, 041K0ZN, 047Y0ZZ, 041M09M, 047T0ZZ, 047Q0ZZ, 041C0JH, 047Q3DZ, 047U3DZ, 047F3ZZ, 041N0JQ, 047U0ZZ, 041H0JH, 047F34Z, 041L0AH, 041J0JJ, 04BM0ZZ, 041M09P, 041L0JQ, 04700DZ, 041J0JH, 04CW3ZZ, 041L4JL, 047E34Z, 03160JB, 04BY0ZZ, 047R3DZ, 041M0JQ, 35637, 04CM4ZZ, 04LL3ZZ, 041L0JS, 04CV3ZZ, 041M09L, 041D0JJ, 041M0JL, 047E3DZ, 04LK3DZ, 04LL0ZZ, 041K0ZM, 041L0KH, 35540, 04CE0ZZ, 04CW0ZZ, 047C4DZ, |

| | | |
|---|---|---|
| | | 061M09Y, 041K4ZL, 041N09P, 041L0ZJ, 04100JG, 35570, 04CE3ZZ, 04CR4ZZ, 041L0AL, 047N0DZ, 047M04Z, 35565, 045Y0ZZ, 047C44Z, 04100JH, 047N4ZZ, 041L0KM, 047N04Z, 03160Z7, 041K0KS, 041K49L, 04CN4ZZ, 041K49N, 041M0JM, 047J44Z, 35879, 047E0DZ, 03150A8, 041L09P, 047D4ZZ, 047M4ZZ, 04CT4ZZ, 04BY4ZZ, 047K44Z, 047034Z, 041M0KS, 041K0KM, 041K0ZS, 047P04Z, 047F0DZ, 047D4DZ, 041N0JL, 04CV0ZZ, 041L4JH, 041N09M, 041L4KL, 041L09S, 041K0JS, 041L0KJ, 047Q4ZZ, 04CP4ZZ, 041K4JJ, 047S4ZZ, 04LN0ZZ, 047C4ZZ, 041L0JK, 041N09L, 03150JC, 041K0KK, 047Y0DZ, 04LU0ZZ, 041K0KJ, 047T4ZZ, 041N0KQ, 35361, 041K0KQ, 04LQ0ZZ, 041L4AL, 04BP0ZZ, 04100KH, 04100ZF, 041L0ZK, 04RK07Z, 041K4JM, 03160A8, 04LK0CZ, 047T0DZ, 04BN0ZZ, 041L4JJ, 041K0AK, 047U0DZ, 041K0AL, 03150Z6, 047N4DZ, 041L0ZS, 04100JR, 041K0ZH, 041N0ZS, 041K09J, 04BS0ZZ, 041L4ZH, 041F0JJ, 041K09P, 041L0AJ, 041L0KQ, 04BR0ZZ, 047J4DZ, 047L4ZZ, 04LR3DZ, 041M0KQ, 041N0KP, 35663, 04CF0ZZ, 04100J7, 04100JQ, 041K4JL, 047E0ZZ, 047U4ZZ, 041L0KS, 041K4JK, 047R44Z, 03150AC, 04LM3ZZ, 03150J7, 04CF3ZZ, 047D44Z, 35884, 04RK0JZ, 047K4ZZ, 03160A7, 04CU4ZZ, 04100JF, 041K0AJ, 04LL0CZ, 047H4ZZ, 041K0AN, 35306, 041K4KN, 04RK0KZ, 041E0JH, 04104JC, 041K4ZM, 047P0DZ, 03150A6, 04RL0KZ, 047H4DZ, 047Q0DZ, 047N44Z, 041N0JP, 041K09H, 04104KD, 04100J6, 041K0KH, 04LK0DZ, 04LP0ZZ, 04100Z8, 04LS3ZZ, 041K4JN, 04104ZK, 041N0ZM, 35881, 04100ZD, 047L4DZ, 041L49L, 047E04Z, 061G0JY, 041D0JH, 041L0AN, 04LR3ZZ, 041M0ZQ, 04100AK, 041N09S, 041N0JM, 047J4ZZ, 041K0ZK, 047H44Z |
